# Genomic epidemiology of SARS-CoV-2 within households in coastal Kenya: a case ascertained cohort study

**DOI:** 10.1101/2022.10.26.22281455

**Authors:** Charles N. Agoti, Katherine E. Gallagher, Joyce Nyiro, Arnold W. Lambisia, Nickson Murunga, Khadija Said Mohammed, Leonard Ndwiga, John M. Morobe, Maureen W. Mburu, Edidah M. Ongera, Timothy O. Makori, My V.T. Phan, Matthew Cotten, Lynette Isabella Ochola-Oyier, Simon Dellicour, Philip Bejon, George Githinji, D. James Nokes

## Abstract

Analysis of severe acute respiratory syndrome coronavirus 2 (SARS-CoV-2) genomic sequences from household infections may provide useful epidemiological information for future control measures. Between December 2020 and July 2022, we conducted a case-ascertained household cohort study whereby households were recruited if a member was either a SARS-CoV-2 case or contact of a confirmed case. A total of 765 members of 214 households were prospectively monitored for SARS-CoV-2 infection and transmission. Follow-up visits collected a nasopharyngeal/oropharyngeal (NP/OP) swab on days 1, 4 and 7 for RT-PCR diagnosis. If any of these were positive, further swabs were collected on days 10, 14, 21 and 28. Of 2,780 NP/OP swabs collected, 540 (19.4%) tested SARS-CoV-2 positive and viral genome sequences were recovered for 288 (53.3%) positive samples. The genomes belonged to 23 different Pango lineages. Phylogenetic analysis including contemporaneous Coastal Kenya data estimated 233 putative transmission events involving 162 members of the 89 households, of which 60 (25%) were intra-household transmission events while 173 (75%) were infections that likely occurred outside the households. In 34 (38%) households, multiple virus introductions were observed (up to six) within the one-month follow-up period, in contrast to high-income settings, where a single introduction seemed to occur during epidemic waves. Our findings suggests that in this setting control of respiratory virus spread by household member isolation will be ineffective.

## Introduction

Households are a fundamental unit of social structure and the frequent scene of respiratory pathogen transmission including for severe acute respiratory syndrome coronavirus 2 (SARS-CoV-2), the aetiological agent of coronavirus disease 2019 (COVID-19) ^1,2^. The household secondary attack rate (i.e., the probability of infection of susceptible household members directly from the first case in the household) for SARS-CoV-2 has been estimated to be about 21.1% (95%CI: 17.4-24.8%) with considerable heterogeneity observed over geographic regions and time periods ^3-6^. An improved understanding of SARS-CoV-2 household transmission, including the frequency of virus transmission within a household compared to transmission from outside into the household, may help refine local control measures.

As of August 2022, Kenya had experienced six waves of SARS-CoV-2 infections ^7^. In this period, Kenya’s COVID-19 countermeasures as measured by the Oxford Stringency Index (SI), a measure based on nine key indicators rescaled from 0-100, fluctuated between 36 and 89^8^ in response to the perceived threat. The different infection waves were dominated by distinct virus variants: the first two by the early ancestral virus lineage (mainly B.1), the third by the Alpha (B.1.1.7) variant of concern (VOC), the fourth by Delta VOC (B.1.617.2), the fifth by Omicron VOC (BA.1), and the sixth by Omicron sub-variants BA.4/5 ^9^. Local epidemiological studies suggest many SARS-CoV-2 infections have been asymptomatic or mild ^10^, as evidenced by the high seroprevalence (48.5% by March 2021 ^11^) without high numbers of hospitalisations or excess mortality ^12^. However, confirmed active SARS-CoV-2 infections by the Kenyan Ministry of Health (MoH) have been reported in less than 1% of the Kenyan population indicating inadequate testing. Consequently, gaps exist in our quantitative documentation of SARS-CoV-2 circulation within the Kenyan population.

To date, SARS-CoV-2 genomic analysis has played a key role in elucidating COVID-19 pandemic transmission patterns ^13-17^. For instance, genomic analysis has helped uncover the nature of virus seeding into close living environments like, hospitals ^18,19^, prisons^20^, cruise ships ^21^, long-term care facilities ^22^, or learning institutions ^23^, and has also uncovered a number of superspreading events ^19,24^. Unlike many RNA viruses, SARS-CoV-2 replication is believed to be under some level of proof-reading^25^, limiting its substitution rate (estimated at 9.90 × 10^−4^ nucleotide substitutions/site/year) ^26^, a parameter critical in applying genomics to understand short-term epidemiological dynamics.

Few studies have examined SARS-CoV-2 genomic diversity in putative within-household transmission cases. In Ireland, Hare *et al* ^27^ found that most family members testing positive had indistinguishable consensus genome sequences from other family members and the early presumed index case had a divergent sub-lineage. In the present study, we document SARS-CoV-2 transmission patterns within households in coastal Kenya by analysing infections identified in a case-ascertained cohort (i.e. households enrolled if a member was confirmed case of SARS-CoV-2 or a contact of one) during successive local waves of infections ^28^. We undertook detailed genomic analyses to identify patterns of SARS-CoV-2 introductions into households and to document the frequency and patterns of infection spread within households in coastal Kenya.

## Results

### Baseline characteristics

A total of 765 participants from 214 households were recruited between 10^th^ December 2020 and 29^th^ July 2022. From these, 2,780 nasopharyngeal/oropharyngeal (NP/OP) swabs were collected, 540 (19.4%) of which tested SARS-CoV-2 positive by RT-PCR (**Table 1**). The positive swabs were from 254 infected participants in 119 households. The temporal distribution of the swab collections for all the 214 households and their RT-PCR testing results are shown in **S1 Fig**. The participants with a positive swab had a median age of 27 years (IQR: 12.0-46.0; **Table 1)**, with 164 (64.6%) females.

**Table 1.** Demographic characteristics of study participants.

| **Characteristic** | **Positive** | **Negative** | **Total** |
| --- | --- | --- | --- |
| **NP/OP swabs** | 540 | 2240 | 2780 |
| **Recruited households** | 119 | 95 | 214 |
| **Recruited participants** | 254 | 511 | 765 |
| **Sex** |  |  |  |
| female (%) | 164 (64.6) | 297 (58.1) | 461 (60.3) |
| male (%) | 90 (35.4) | 214 (41.9) | 304 (39.7) |
| **Age distribution (years)** |  |  |  |
| Median (IQR) | 27.0 (12.0-46.0) | 20.0 (10.0-40.0) | 24.0 (11.0-44.0) |
| **Age categories** |  |  |  |
| 0-4 y (%) | 31 (12.2) | 54 (10.6) | 85 (11.1) |
| 5-9 y (%) | 23 (9.1) | 54 (10.6) | 77 (10.1) |
| 10-19 y (%) | 48 (18.9) | 133 (26.0) | 181 (23.7) |
| 20-39 y (%) | 73 (28.7) | 113 (28.6) | 219 (28.6) |
| 40-64 y (%) | 63 (24.8) | 92 (18.0) | 155 (20.3) |
| 65+ y (%) | 14 (5.5) | 31 (6.1) | 45 (5.9) |
| Unknown | 2 (0.8) | 1 (0.2) | 3 (0.4) |
| **Symptom status** |  |  |  |
| asymptomatic | 82 (32.3) | 345 (67.5) | 427 (55.8) |
| symptomatic | 172 (67.7) | 166 (32.5) | 338 (44.2) |

IQR: Interquartile Range, y: years

Compared to participants who remained SARS-CoV-2 negative during the follow-up period, positive cases were more likely to report at least one acute respiratory illness (ARI) symptom (67.7% vs 32.5%; *p <0*.*001*, chi-squared test). The household recruitments were predominantly coincidental with the national COVID-19 waves 3, 4, 5 and 6 (**Fig. 1A and B**) with only one household recruited during wave 2. The changes in the national countermeasures during the study period as estimated by the Oxford SI is provided in **Fig. 1C**.

**Fig. 1.**
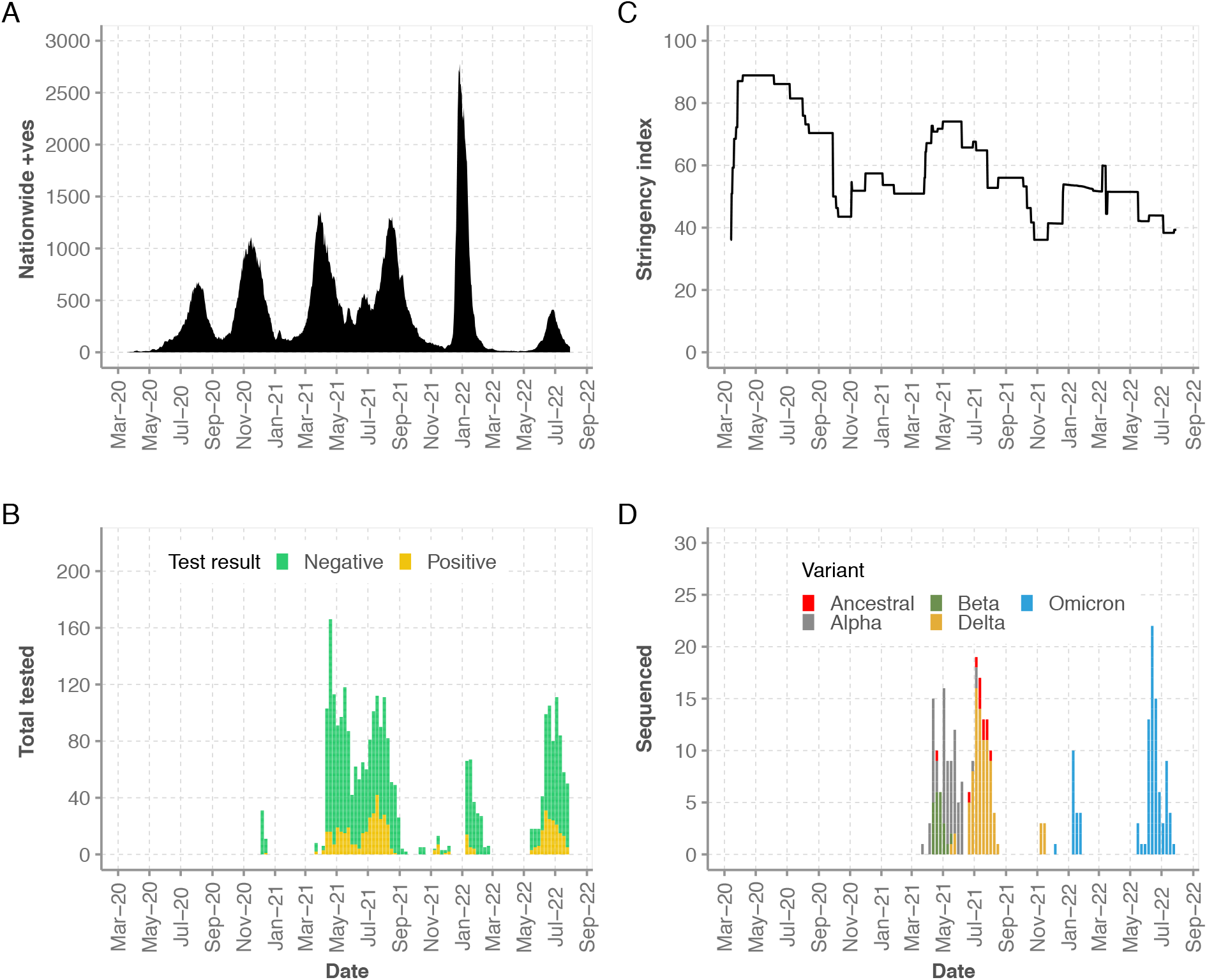
The timeline of the household study and genomic sequencing results. Panel A: Reported SARS-CoV-2 infections observed in Kenya between March 2020 and March 2022. Panel B: Temporal distribution of the collected NP/OPs and their RT-PCR diagnostic results. Panel C: The level of government restrictions. Panel D: Temporal distribution of the SARS-CoV-2 VOCs detected in the household study.

### Genomic sequencing and lineage/VOC classification

Near complete genomes (over 80% coverage) were obtained from 288 samples (53.3% of positive samples) collected from 162 participants from 89 households. The samples that failed sequencing (n = 252) had high Ct values (corresponding to low viral load; >33.0; **S2. Fig**.) or yielded low quality PCR products during library preparation. The 288 genomes were classified into non-VOC (Pango lineage B.1; n = 11), Alpha VOC (n = 70), Beta VOC (n = 22), Delta VOC (n = 88) and Omicron VOC (n = 97; **Fig.1D**). All Alpha and Beta sequences fell within lineage B.1.1.7 and B.1.351, respectively. Within the Delta VOC, six Pango lineages were identified while within the Omicron VOC 14 Pango lineages were identified. The lineages within Omicron VOC included those that classified under BA.1, BA.2, BA.4 and BA.5 sub-variants. A summary of the temporal distribution of the 23 Pango lineages that were identified across the sequenced cases and their history of global detection is presented in **S2 Fig. and S1 Table**.

### Phylogenetic clustering of the household study genomes

To investigate the genetic diversity in the household study genome sequences, we reconstructed a maximum likelihood (ML) phylogeny that included background coastal Kenya co-circulating viruses (n = 2,555). As expected, the household genome sequences clustered by VOC and Pango lineages with other Kenyan coastal sequences (**S3 Fig.)**. Notably, lineage B.1 sequences were found in multiple branches of the phylogeny, including some at the base of branches leading to Beta and Delta VOCs. To assess the genetic relatedness of the recovered genomes within and between households, we reconstructed VOC-specific phylogenies with tips coloured by the household of sampling (**Fig. 2**). Here we observed both intra- and inter-household clustering (i.e., multiple sequences of a single clade identified in one household or shared between two or more households). In a few households, sequences of a single VOC but distinct clades were observed, indicating multiple distinct virus introductions into the same household (**Fig. 2**).

**Fig. 2.**
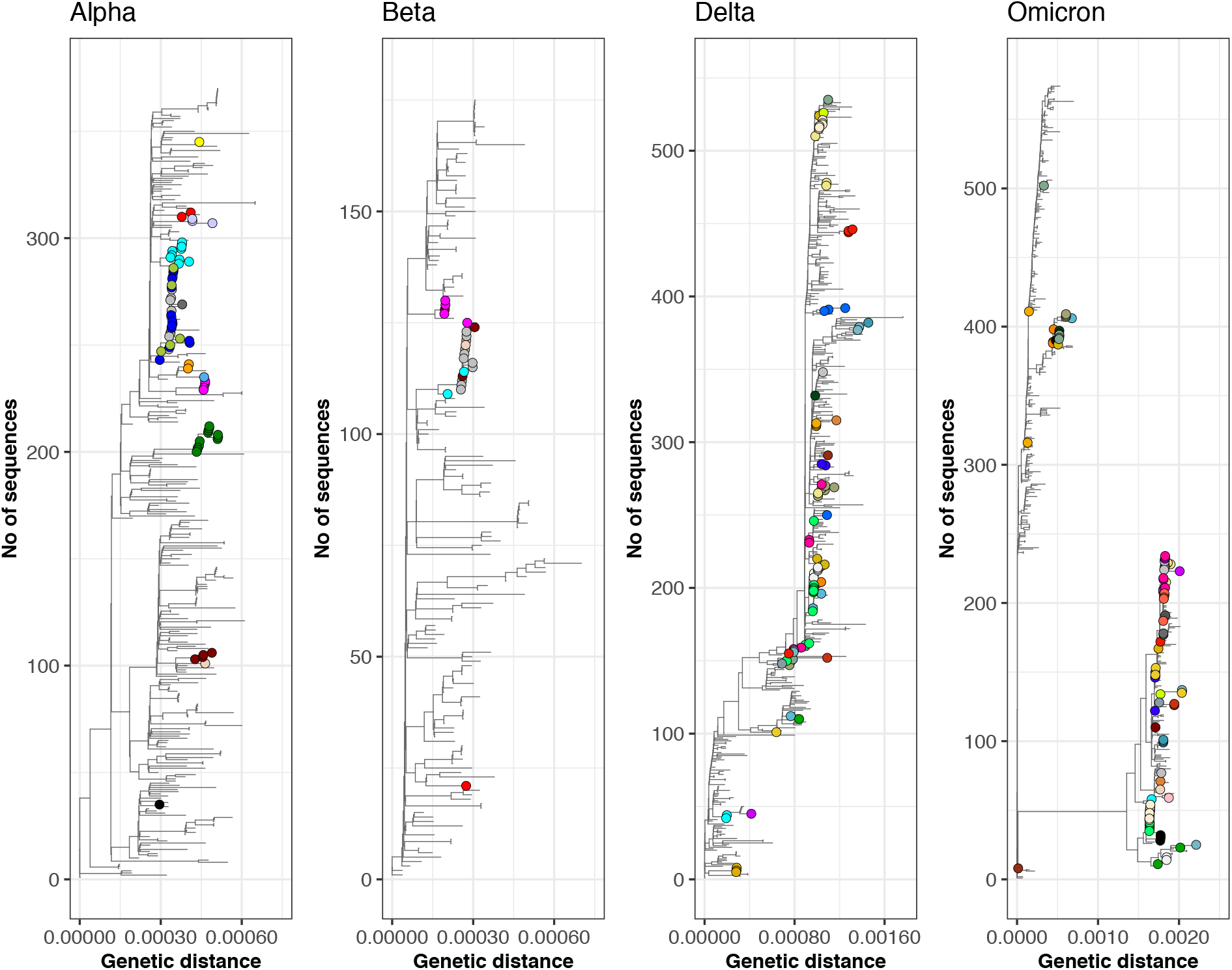
Phylogenetic patterns of variants of concern (VOC) in the household study using maximum-likelihood methods. These household study VOC phylogenies include other SARS-CoV-2 genome sequences generated from samples collected in six Kenyan coastal counties (Mombasa, Kwale, Kilifi, Taita Taveta, Tana River and Lamu) during the study period as background diversity (tips without symbols). On the phylogenies, household sample derived sequences are displayed as filled circles, coloured distinctly by household. In the Alpha, Beta, Delta, and Omicron phylogenies, 370, 175, 535 and 574 genome sequences were included, respectively.

### Estimating the number of introductions into the households

SARS-CoV-2 has been reported to have a genomic evolutionary rate of ∼2 nucleotide substitutions per genome per month ^29^. A heterogenous distribution of the pairwise nucleotide differences of specimens identified in the same household was observed (range 0-63; median 0.0, mean 2.8; **S4 Fig**.). More than two nucleotide differences between genomes from the same household were observed in 25 households, implying possible multiple introductions into these households. We further investigated the number of virus introductions into each household using ancestral state reconstruction (ASR) along the dated ML phylogeny ^16,30^. A total of 155 virus introductions were predicted into the 89 households where we recovered SARS-CoV-2 sequence data. On classifying the transition events by sequence origin (“non-household” events - those individuals not from same household - and “household” events - those where both involved individuals were members of same household), we found that most transitions were “non-household” compared to “household” transition events (75% vs. 25%; **Fig. 3A**). Overall, we estimated that a single virus introduction occurred for 55 households (61%), two introductions for 16 households (17%), three introductions for nine households (10%), four introductions for five households (6%), five introductions for three households (3%) and six introductions in one household (**Fig. 3B**).

**Fig. 3.**
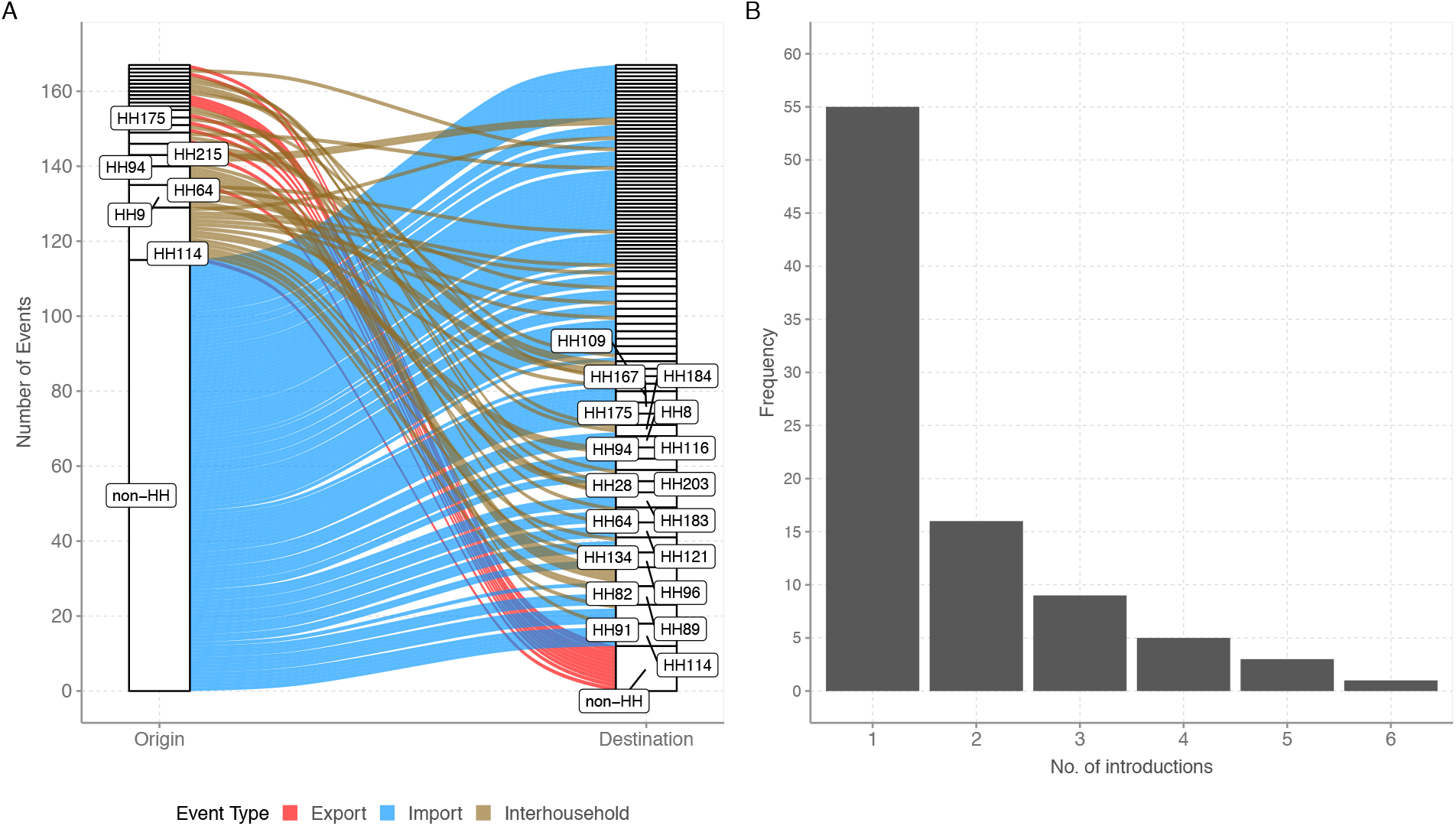
Patterns of SARS-CoV-2 introductions into studied households as determined by ancestral state reconstruction. Panel A: Alluvial plot showing the number of viral imports and exports into and out of study households. Panel B: Frequency of distinct introductions into the study households.

## Discussion

We provide evidence of frequent multiple introductions of SARS-CoV-2 into rural coastal Kenyan households within a month, a finding that was unexpected. The conventional view has been that households with concurrently infected members most likely acquire the infection introduced through a single index case. This assumption has been repeatedly supported by several genomic studies. For instance, a phylogenetic analysis performed in a Dutch study following 85 households showed a single introduction into all the studied households ^4^. By contrast, in the present study, about one-third of infected households (38%) had two or more introduction events within a distinct epidemic wave.

A variety of factors may explain the differences in household SARS-CoV-2 introduction patterns identified in our study compared to previous observations. First, in our setting, multiple families may live in one compound and eat from one kitchen and these were defined as a single household in this study ^31^. Such living arrangement results in relatively larger household sizes that may be associated with higher chances of multiple virus introductions, especially at the peak of epidemic waves. Second, the dominance of informal sector jobs in this setting may have made options like working from home difficult to implement. As result the potential for acquiring infection outside the household setting was high relative to high-income settings.

Our study followed-up participants for a period of up to 1 month with serial sample collection and recovered genomes were analysed in the context of contemporaneous locally circulating diversity in coastal Kenya ^32^. Despite observing minimal nucleotide variation between samples from members of the same household infection clusters, when we incorporated sampling dates through the ASR analysis, we were able to partially reconstruct potential within-household transmission events. This allowed the identification of multiple introductions into the households of closely related viruses, observation of putative within-household transmission and short-interval reinfections.

Few studies have examined SARS-CoV-2 households transmission dynamics within Africa ^33-35^, and these have resulted in diverse findings. In rural Egypt, a 6-month study reported a SAR of 89.8% ^33^, in South Africa a 13-month study reported a 25% infection rate among vulnerable household contacts ^34^, and in Madagascar, a SAR of 38.8% (CI:19.5-57.2) ^36^ was reported. None of these studies included a viral genomic analysis to support any conclusions that the inferred household transmission clusters were epidemiologically linked and arose from a single index case.

The Kenyan government countermeasures in place during the study period may have had an impact on the spread of SARS-CoV-2 within the study households. In June 2020, the Kenyan government announced guidelines for home-based care for asymptomatic or mildly symptomatic patients without co-morbidities ^37^. Kenya started vaccinating its population in March 2021, but the coverage was low (<15%) during the study period ^38^, and thus it is unlikely that it affected transmission during our study. The stringency index in the country during the study period fluctuated from 35% to 75%. However, we did not detect variation in the pattern of introductions over time, which could suggest that the various restrictions had minimal impact at the household level. However, concluding on this would likely require more advanced epidemiological modelling.

This study had several limitations. First, several positive NP/OP samples (46.7%) failed to yield viral genome sequences or had large genome sequencing gaps due to PCR amplicon drop-off. With such a high level of genome data missingness, the overall phylogenetic signal was reduced in trying to establish who infected whom or directionality of transmission. Second, we cannot rule out that a few of the sequence changes observed could be sequencing or assembly artifacts ^39^. Third, the case-ascertained study design we used had the drawback that by the time the first sample was collected, multiple positive cases had already occurred in most households. Most of the index cases were recruited following presentation to a health facility with ARI. This complicated our effort of fully determining who infected whom back in the household. To overcome this challenge, future studies should observe members before entry of the virus into households and genomic data co-analysed with other relevant epidemiological data ^40^. Fourth, our sampling interval, especially after week two, may have missed persons who had been positive for less than the 7 days sample collection interval. Higher density sampling has previously been associated with a higher attack rate^4^. Finally, we did not bring in other datatypes like, contact patterns, symptoms and shared intra-host variation ^41^, which would have provided further insights into potential transmission linkages. An analysis of secondary attack rates is being conducted that includes all data, including serological evidence of infection, which is not included here.

In conclusion, we identified an unusually high number of independent virus introductions into households in coastal Kenya during household temporally clustered infections. Our findings suggests that control of SARS-CoV-2 transmission by household member isolation alone may not stop community transmission in this setting. Our study further highlights the importance of examining genomic data for accurate estimation and interpretation of SARS-CoV-2 household epidemiological parameters.

## Methods

### Study design and recruitment

We conducted a case-ascertained study in coastal Kenya, where new households were recruited into the study via five local health facilities or the Kilifi County Department of Health rapid response team (RRTs). Households were defined as dwellings or groups of dwellings that share the same kitchen or cooking space (**S2 Table**). Many of the recruited households were from within the Kilifi Health and Demographic Surveillance System (KHDSS) area located in Kilifi County, Coastal Kenya ^42^. To get enrolled, a household needed to have at least two occupants, to be accessible by road, and to obtain permission from the household head. In the beginning, only households with a member who was a contact of a confirmed case from a different household were recruited, but due to slow enrolment, we revised the protocol to include households with confirmed cases at first sampling. A household was exempted if at the time of recruitment: two or more members had already developed COVID-19 symptoms (e.g. fever, sore throat, cough etc), a member had been hospitalized due to COVID-19, or the household had been enrolled in a trial of therapeutic COVID-19 product.

### Follow-up

During each household visit, a NP/OP swab was obtained from all participants and transported in cool boxes with ice packs to KEMRI-Wellcome Trust Research Programme (KWTRP) laboratories within 48 hr for real-time RT-PCR testing. The study had broadly two follow-up arms: “reduced follow-up” and “intense follow-up”. Households in the “reduced follow-up” arm were those where all the members tested SARS-CoV-2 negative at day 1, 4, and 7. Therefore, in that case, follow-up was discontinued henceforth with a few exceptions (**S1 Fig**.). The “intense follow-up arm” was activated when a household member tested positive on day 1, 4, or 7. In that case, the household was sampled again on day 10, 14, 21, and 28. Data on baseline household and demographic characteristics were collected by the study team at enrolment. During all households’ visits, data on presence of ARI symptoms (e.g., fever, cough, runny nose, sore throat, headache) were collected.

### Laboratory procedures

#### SARS-CoV-2 diagnosis

SARS-CoV-2 testing of study samples was undertaken alongside samples collected in six coastal counties of Kenya as part of the national COVID-19 testing as previously described ^10^. Four different viral RNA extraction kits (namely, QIAamp Viral RNA Mini Kit, RNeasy ® QIAcube ® HT Kit, TIANamp Virus RNA Kit and Da An Gene Nucleic acid Isolation and Purification Kit) were deployed in combination with five different RT-PCR kits/protocol (namely, Da An Gene Co. detection Kit, European Virus Archive-Global (EVAg) E gene protocol, Standard M Kit, Sansure Biotech Novel Coronavirus (2019-nCoV) Nucleic Acid Diagnostic Real-time RT-PCR kit^10^). Positives were determined using the kit/protocol-defined cycle thresholds (Ct). In kits where multiple SARS-CoV-2 genomic regions were targeted, the average cycle threshold (Ct) was calculated from the individual Cts.

#### Genome sequencing

We aimed to sequence whole genomes of all the RT-PCR positive samples with a cycle threshold of < 33.0. Viral RNA was re-extracted from the specimens using QIAamp viral RNA mini-Kit following the manufacturer’s instructions and converted to cDNA using Lunascript kit with ARTIC protocol primers ^43^. Genome amplification was conducted using Q5 PCR kit and ARTIC protocol primers (initially v3 and then v4). Sequencing libraries were prepared using Oxford Nanopore Technologies (ONT) ligation sequencing kit SQK-LSK109 and the ONT Native Barcoding Expansion kit as described in the ARTIC protocol ^43^. Sequencing was performed on Oxford Nanopore Technologies’ MinION or GridION devices using R9.4.1 flow cells.

### Bioinformatic analysis

#### Genome assembly and lineage assignment

The ONT’s raw sequencing reads (FAST5) were base-called and demultiplexed using ONT’s Guppy v3.5-4.2. The resultant files (FASTQ) were assembled into consensus genomes using ARTIC bioinformatic pipeline reference-based approach (https://artic.network/ncov-2019/ncov2019-bioinformatics-sop.html; last accessed 2022-09-17). Only nucleotides with a read depth of more than × 20 were included into the consensus sequence. Only sequences with >80% genome coverage were further analysed. The genomes were assigned Pango lineages using the command-line installation of pangolin v4.1.3, PUSHER-v1.3, scorpio v0.3.16 and constellation v0.1.6 ^44,45^.

#### Phylogenetic analysis

Multiple sequence alignments were generated using Nextalign v.1.10.1 referenced-based aligner within the Nextclade tool v0.14.2 ^46^ using the command:

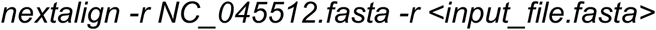

Alignments were visualized using a custom Python script and “snipit” tool (https://github.com/aineniamh/snipit; last accessed 2022-05-20). Pairwise distances were calculated using pairsnp.py (https://github.com/gtonkinhill/pairsnp/; last accessed 2022-05-20) using the command:

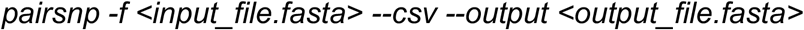

Phylogenetic relationships between all recovered genomes and between viruses classified under the same VOC were obtained through the inference of maximum likelihood (ML) trees performed with the program IQTREE v2.1.3 with a general time reversible (GTR) substitution model using the command:

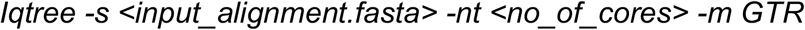

We included contemporaneous genomes from the six coastal Kenya counties (Mombasa, Kilifi, Kwale, Taita Taveta, Tana River, and Lamu) that were sequenced as part of the national SARS-CoV-2 genomic surveillance to provide phylogenetic context to the household study genomes. Each ML tree was subsequently time-calibrated using the program TreeTime, assuming a constant evolutionary rate of SARS-CoV-2 genome of 8.0 × 10^−4^ using the command:

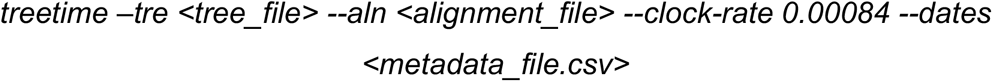

The phylogenetic trees were combined with metadata and visualized with the R package “ggtree” v2.4.2 ^47^.

#### Virus introductions and putative transmissions

The number of independent virus introductions into the households was investigated using two approaches.

##### i. Pairwise nucleotide substitution

Here, we compared observed nucleotide differences between pairs of household genomes with the number of nucleotide mutations expected over the time interval between the two sampling dates. The pairwise differences between the household study genomes were computed using the program pairsnp (https://github.com/gtonkinhill/pairsnp; accessed 06-Feb-2023).

##### ii. Ancestral state reconstruction (ASR)

ASR approach was used to identify the introduction events into a household and count the transitions between household members ^16^. This was performed along the time-scaled phylogeny obtained for each VOC. To infer the number of introduction events into each household, a variable “source” was generated for which the sequences were assigned the household ID or noted as “non-household”. Using the dated phylogeny, the nucleotide sequence alignment, the “source” metadata file, a mugration analysis was run using TreeTime^48^, using the command:

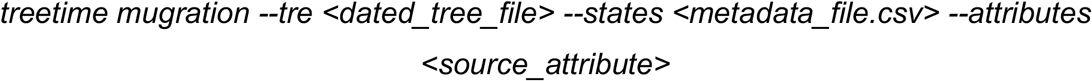

To infer within household transition events, a variable “participant” was generated for which the sequences were assigned the participant ID or noted as “non-household”. The mugration analysis was rerun using the dated phylogeny and transitions between members of the same households noted using python scripts described in ^16,30^.

### Statistical analysis

Summary statistics were computed for key demographic characteristics including mean, median, standard deviation as appropriate. Infection prevalence was expressed using proportions and comparison between groups included appropriate statistical tests (e.g., chi-square or Fisher’s exact). All statistical analyses were performed in R version 4.0.5.

### Ethical consideration

The study protocol was reviewed and approved by both the Scientific and Ethics Research Unit (SERU) at Kenya Medical Research Institute (KEMRI), Nairobi, Kenya (REF: SERU protocol # 4077) and the University of Warwick, Biomedical and Scientific Research Ethics Committee, Coventry, United Kingdom (REF: BSREC 150/19-20 AM01). Prior to baseline data and biospecimen collection, written informed consent was obtained from all participants aged 18 years or older, while for participants aged less than 18 years consent was obtained from their parents or legal guardians. Assent was also sought for adolescents (13-17 years of age).

## Data Availability

The consensus genome sequences obtained in this study that passed our quality control filters have been submitted to both GISAID and GenBank database (accession numbers available in appendix pages of the supplementary material). The raw data files have been prepared for deposition in Harvard DataVerse. For more detailed information beyond the metadata used in the paper, there is a process of managed access requiring submission of a request form for consideration by our Data Governance Committee (http://kemri-wellcome.org/about-us/#ChildVerticalTab_15).

## Code availability

The code for the analyses presented in this manuscript is available from Github using the link https://github.com/cnyaigoti/Household_study_2021-2022.

## Acknowledgements

We thank (a) the members of the Kilifi County rapid response team who worked with our field study team in collecting the samples analysed here; (b) the members of the COVID-19 KWTRP Testing Team who undertook real-time RT-PCR processing of the samples received at KWTRP to identify positives (see full list of members below). This work was supported by the National Institute for Health and Care Research (NIHR) (project reference 17/63/82) using UK aid from the UK Government to support global health research, The UK Foreign, Commonwealth and Development Office and Wellcome Trust (grant# 220985). Members of COVID-19 Testing Team at KWTRP are supported by multiple funding sources including UNITAD (BOHEMIA study received by Dr Marta Maia funded UNITAID), EDCTP (Senior Fellowship and Research and Innovation Action (RIA) grants received by Dr Francis Ndungu), GAVI (PCIVS grant received by Prof. Anthony Scott). Dr Simon Dellicour acknowledges support from the *Fonds National de la Recherche Scientifique* (F.R.S.-FNRS, Belgium; grant n°F.4515.22), from the Research Foundation - Flanders (*Fonds voor Wetenschappelijk Onderzoek-Vlaanderen*, FWO, Belgium; grant n°G098321N), and from the European Union Horizon 2020 project MOOD (grant agreement n°874850). MP and MC were supported by the Wellcome Trust and FCDO - Wellcome Epidemic Preparedness – Coronavirus (AFRICO19, grant agreement number 220977/Z/20/Z). The views expressed in this publication are those of the author (s) and not necessarily those of NIHR, the Department of Health and Social Care, Foreign Commonwealth and Development Office, Wellcome Trust or the UK government.

## List of members of the COVID-19 Testing Team at KWTRP

Agnes Mutiso, Alfred Mwanzu, Angela Karani, Bonface M. Gichuki, Boniface Kaaria, Brian Bartilol, Brian Tawa, Calleb Odundo, Caroline Ngetsa, Clement Lewa, Daisy Mugo, David Amadi, David Ireri, Debra Riako, Domtila Kimani, Donwilliams Omuoyo, Edwin Machanja, Elijah Gicheru, Elisha Omer, Faith Gambo, Horace Gumba, Isaac Musungu, James Chemweno, Janet Thoya, Jedida Mwacharo, Jennifer Musyoki, John Gitonga, Johnstone Makale, Justine Getonto, Kelly Ominde, Kelvias Keter, Lydia Nyamako, Margaret Nunah, Martin Mutunga, Metrine Tendwa, Moses Mosobo, Nelson Ouma, Nicole Achieng, Patience Kiyuka, Perpetual Wanjiku, Peter Mwaura, Rita Warui, Robinson Cheruiyot, Salim Mwarumba, Shaban Mwangi, Shadrack Mutua, Susan Njuguna, Victor Osoti, Wesley Cheruiyot, Wilfred Nyamu, Wilson Gumbi and Yiakon Sein.

## Author’s contributions

The project was conceived and designed by CNA, KEG, JUN, MC, and DJN; Laboratory processing of specimens was conducted by JUN, LIO, NM, AWL, KSM, LN, JMM, MWM, EMO, and TOM; Management and analysis of data were handled by AWL, CNA, NM, SD and GG. CNA wrote the first draft; MP, MC, LIO, SD, PB, and DJN critically reviewed the manuscript to produce the final draft.

## References

1 Lee, E. C., Wada, N. I., Grabowski, M. K., Gurley, E. S. & Lessler, J. The engines of SARS-CoV-2 spread. Science 370, 406–407 (2020). https://doi.org:doi:10.1126/science.abd8755

2 Park, Y. J. et al. Contact Tracing during Coronavirus Disease Outbreak, South Korea, 2020. Emerging Infectious Disease journal 26, 2465 (2020). https://doi.org:10.3201/eid2610.201315

3 Thompson, H. A. et al. Severe Acute Respiratory Syndrome Coronavirus 2 (SARS-CoV-2) Setting-specific Transmission Rates: A Systematic Review and Meta-analysis. Clin Infect Dis 73, e754–e764 (2021). https://doi.org:10.1093/cid/ciab100

4 Kolodziej, L. M. et al. High SARS-CoV-2 household transmission rates detected by dense saliva sampling. Clinical Infectious Diseases (2022). https://doi.org:10.1093/cid/ciac261

5 Madewell, Z. J., Yang, Y., Longini, I. M., Jr, Halloran, M. E. & Dean, N. E. Household Secondary Attack Rates of SARS-CoV-2 by Variant and Vaccination Status: An Updated Systematic Review and Meta-analysis. JAMA Network Open 5, e229317–e229317 (2022). https://doi.org:10.1001/jamanetworkopen.2022.9317

6 Jørgensen, S. B., Nygård, K., Kacelnik, O. & Telle, K. Secondary Attack Rates for Omicron and Delta Variants of SARS-CoV-2 in Norwegian Households. JAMA 327, 1610–1611 (2022). https://doi.org:10.1001/jama.2022.3780

7 Tegally, H. et al. The evolving SARS-CoV-2 epidemic in Africa: Insights from rapidly expanding genomic surveillance. Science, eabq5358 (2022). https://doi.org:10.1126/science.abq5358

8 Hale, T. et al. A global panel database of pandemic policies (Oxford COVID-19 Government Response Tracker). Nat Hum Behav 5, 529–538 (2021). https://doi.org:10.1038/s41562-021-01079-8

9 Nasimiyu, C. et al. Imported SARS-CoV-2 Variants of Concern Drove Spread of Infections across Kenya during the Second Year of the Pandemic. COVID 2, 586–598 (2022).

10 Nyagwange, J. et al. Epidemiology of COVID-19 infections on routine polymerase chain reaction (PCR) and serology testing in Coastal Kenya. Wellcome Open Res 7, 69 (2022). https://doi.org:10.12688/wellcomeopenres.17661.1

11 Uyoga, S. et al. Prevalence of SARS-CoV-2 Antibodies From a National Serosurveillance of Kenyan Blood Donors, January-March 2021. Jama 326, 1436–1438 (2021). https://doi.org:10.1001/jama.2021.15265

12 Otiende, M. et al. Impact of COVID-19 on mortality in coastal Kenya: a longitudinal open cohort study. medRxiv, 2022.2010.2012.22281019 (2022). https://doi.org:10.1101/2022.10.12.22281019

13 Li, J., Lai, S., Gao, G. F. & Shi, W. The emergence, genomic diversity and global spread of SARS-CoV-2. Nature 600, 408–418 (2021). https://doi.org:10.1038/s41586-021-04188-6

14 Bugembe, D. L. et al. Main Routes of Entry and Genomic Diversity of SARS-CoV-2, Uganda. Emerg Infect Dis 26, 2411–2415 (2020). https://doi.org:10.3201/eid2610.202575

15 Githinji, G. et al. Tracking the introduction and spread of SARS-CoV-2 in coastal Kenya. Nat Commun 12, 4809 (2021). https://doi.org:10.1038/s41467-021-25137-x

16 Wilkinson, E. et al. A year of genomic surveillance reveals how the SARS-CoV-2 pandemic unfolded in Africa. Science 374, 423–431 (2021). https://doi.org:10.1126/science.abj4336

17 Vöhringer, H. S. et al. Genomic reconstruction of the SARS-CoV-2 epidemic in England. Nature 600, 506–511 (2021). https://doi.org:10.1038/s41586-021-04069-y

18 Ellingford, J. M. et al. Genomic and healthcare dynamics of nosocomial SARS-CoV-2 transmission. Elife 10 (2021). https://doi.org:10.7554/eLife.65453

19 Illingworth, C. J. et al. Superspreaders drive the largest outbreaks of hospital onset COVID-19 infections. Elife 10 (2021). https://doi.org:10.7554/eLife.67308

20 Hershow, R. B. et al. Rapid Spread of SARS-CoV-2 in a State Prison After Introduction by Newly Transferred Incarcerated Persons - Wisconsin, August 14-October 22, 2020. MMWR Morb Mortal Wkly Rep 70, 478–482 (2021). https://doi.org:10.15585/mmwr.mm7013a4

21 Hoshino, K. et al. Transmission dynamics of SARS-CoV-2 on the Diamond Princess uncovered using viral genome sequence analysis. Gene 779, 145496 (2021). https://doi.org:10.1016/j.gene.2021.145496

22 Aggarwal, D. et al. The role of viral genomics in understanding COVID-19 outbreaks in long-term care facilities. Lancet Microbe 3, e151–e158 (2022). https://doi.org:10.1016/s2666-5247(21)00208-1

23 Baumgarte, S. et al. Investigation of a Limited but Explosive COVID-19 Outbreak in a German Secondary School. Viruses 14 (2022). https://doi.org:10.3390/v14010087

24 Popa, A. et al. Genomic epidemiology of superspreading events in Austria reveals mutational dynamics and transmission properties of SARS-CoV-2. Sci Transl Med 12 (2020). https://doi.org:10.1126/scitranslmed.abe2555

25 Smith, E. C., Blanc, H., Surdel, M. C., Vignuzzi, M. & Denison, M. R. Coronaviruses lacking exoribonuclease activity are susceptible to lethal mutagenesis: evidence for proofreading and potential therapeutics. PLoS Pathog 9, e1003565 (2013). https://doi.org:10.1371/journal.ppat.1003565

26 Nie, Q. et al. Phylogenetic and phylodynamic analyses of SARS-CoV-2. Virus Res 287, 198098 (2020). https://doi.org:10.1016/j.virusres.2020.198098

27 Hare, D. et al. Genomic epidemiological analysis of SARS-CoV-2 household transmission. Access Microbiol 3, 000252 (2021). https://doi.org:10.1099/acmi.0.000252

28 Githinji, G. et al. The genomic epidemiology of SARS-CoV-2 variants of concern in Kenya. medRxiv, 2022.2010.2026.22281446 (2022). https://doi.org:10.1101/2022.10.26.22281446

29 Duchene, S. et al. Temporal signal and the phylodynamic threshold of SARS-CoV-2. Virus Evolution 6 (2020). https://doi.org:10.1093/ve/veaa061

30 Tegally, H. et al. Sixteen novel lineages of SARS-CoV-2 in South Africa. Nat Med 27, 440–446 (2021). https://doi.org:10.1038/s41591-021-01255-3

31 Munywoki, P. K. et al. The source of respiratory syncytial virus infection in infants: a household cohort study in rural Kenya. J Infect Dis 209, 1685–1692 (2014). https://doi.org:10.1093/infdis/jit828

32 Agoti, C. N. et al. Transmission networks of SARS-CoV-2 in Coastal Kenya during the first two waves: a retrospective genomic study. eLife 11, e71703 (2022). https://doi.org:10.7554/eLife.71703

33 Gomaa, M. R. et al. Incidence, household transmission, and neutralizing antibody seroprevalence of Coronavirus Disease 2019 in Egypt: Results of a community-based cohort. PLoS Pathog 17, e1009413 (2021). https://doi.org:10.1371/journal.ppat.1009413

34 Cohen, C. et al. SARS-CoV-2 incidence, transmission, and reinfection in a rural and an urban setting: results of the PHIRST-C cohort study, South Africa, 2020-21. Lancet Infect Dis (2022). https://doi.org:10.1016/s1473-3099(22)00069-x

35 Semakula, M. et al. The secondary transmission pattern of COVID-19 based on contact tracing in Rwanda. BMJ Global Health 6, e004885 (2021). https://doi.org:10.1136/bmjgh-2020-004885

36 Ratovoson, R. et al. Household transmission of COVID-19 among the earliest cases in Antananarivo, Madagascar. Influenza Other Respir Viruses 16, 48–55 (2022). https://doi.org:10.1111/irv.12896

37 Herman-Roloff, A. et al. Adapting Longstanding Public Health Collaborations between Government of Kenya and CDC Kenya in Response to the COVID-19 Pandemic, 2020-2021. Emerg Infect Dis 28, S159–s167 (2022). https://doi.org:10.3201/eid2813.211550

38 Orangi, S. et al. Epidemiological impact and cost-effectiveness analysis of COVID-19 vaccination in Kenya. BMJ Glob Health 7 (2022). https://doi.org:10.1136/bmjgh-2022-009430

39 Bendall, E. E. et al. SARS-CoV-2 Genomic Diversity in Households Highlights the Challenges of Sequence-Based Transmission Inference. mSphere 7, e00400–00422 (2022). https://doi.org:doi:10.1128/msphere.00400-22

40 Agoti, C. N. et al. Genomic analysis of respiratory syncytial virus infections in households and utility in inferring who infects the infant. Sci Rep 9, 10076 (2019). https://doi.org:10.1038/s41598-019-46509-w

41 Gallego-García, P. et al. Limited genomic reconstruction of SARS-CoV-2 transmission history within local epidemiological clusters. Virus Evolution 8 (2022). https://doi.org:10.1093/ve/veac008

42 Scott, J. A. et al. Profile:The Kilifi Health and Demographic Surveillane System (KHDSS). Int J Epidemiol 41, 650–657 (2012). https://doi.org:dys062 [pii] 10.1093/ije/dys062

43 Tyson, J. R. et al. Improvements to the ARTIC multiplex PCR method for SARS-CoV-2 genome sequencing using nanopore. bioRxiv (2020). https://doi.org:10.1101/2020.09.04.283077

44 O’Toole, Á. et al. Assignment of epidemiological lineages in an emerging pandemic using the pangolin tool. Virus Evol 7, veab064 (2021). https://doi.org:10.1093/ve/veab064

45 Rambaut, A. et al. A dynamic nomenclature proposal for SARS-CoV-2 lineages to assist genomic epidemiology. Nat Microbiol 5, 1403–1407 (2020). https://doi.org:10.1038/s41564-020-0770-5

46 Hadfield, J. et al. Nextstrain: real-time tracking of pathogen evolution. Bioinformatics 34, 4121–4123 (2018). https://doi.org:10.1093/bioinformatics/bty407

47 Yu, G., Smith, D. K., Zhu, H., Guan, Y. & Lam, T. T.-Y. ggtree: an r package for visualization and annotation of phylogenetic trees with their covariates and other associated data. Methods in Ecology and Evolution 8, 28–36 (2017). https://doi.org:https://doi.org/10.1111/2041-210X.12628

48 Sagulenko, P., Puller, V. & Neher, R. A. TreeTime: Maximum-likelihood phylodynamic analysis. Virus Evol 4, vex042 (2018). https://doi.org:10.1093/ve/vex042

