## Supplemental file for "Genomic epidemiology of SARS-CoV-2 within households in coastal Kenya: a case ascertained cohort study"

**Affiliations**
[1]Kenya Medical Research Institute (KEMRI)-Wellcome Trust Research Programme (KWTRP), Kilifi, Kenya.
[2]Pwani University, Kilifi, Kenya.
[3]London School of Hygiene & Tropical Medicine, United Kingdom.
[4]Medical Research Centre (MRC)/ Uganda Virus Research Institute, Entebbe, Uganda
[5]MRC-University of Glasgow, Centre for Virus Research Glasgow, United Kingdom.
[6]Spatial Epidemiology Lab (SpELL), Université Libre de Bruxelles, Bruxelles, Belgium.
[7]Department of Microbiology, Immunology and Transplantation, Rega Institute, Laboratory for Clinical and Epidemiological Virology, KU Leuven, University of Leuven, Leuven, Belgium.
[8]Nuffield Department of Medicine, University of Oxford, Oxford, United Kingdom.
[9]School of Life Sciences and Zeeman Institute for Systems Biology and Infectious Disease Epidemiology Research (SBIDER), University of Warwick, Coventry, United Kingdom.

**S1 Table. Lineages identified in the sequenced samples from the present household study. For each lineage we report, their frequency and as well as the global history of their detection .**

| Lineage | Number | VOC status | Most common countries | Earliest date | Description |
|---|---|---|---|---|---|
| B.1 | 12 | non-VOC | United States of America 46.0%, Turkey 12.0%, United Kingdom 7.0%, Canada 4.0%, France 3.0% | 2020-01-01 | A large European lineage the origin of which roughly corresponds to the Northern Italian outbreak early in 2020. |
| B.1.1.7 | 70 | Alpha | United Kingdom 24.0%, United States of America 20.0%, Germany 9.0%, Sweden 6.0%, Denmark 6.0% | 2020-09-01 | UK lineage of concern, associated with the N501Y mutation. More information can be found at cov-lineages.org/global_report.html |
| B.1.351 | 21 | Beta | South_Africa 19.0%, Philippines 9.0%, United States of America 9.0%, Sweden 8.0%, Germany 7.0% | 2020-09-01 | Lineage of concern detected in South Africa |
| AY.122 | 4 | Delta | Germany 17.0%, United States of America 13.0%, France 10.0%, Denmark 10.0%, Sweden 5.0% | 2020-05-11 | Alias of B.1.617.2.122, European lineage |
| AY.4 | 1 | Delta | United Kingdom 84.0%, Denmark 3.0%, Germany 2.0%, Ireland 2.0%, France 1.0% | 2020-05-11 | Alias of B.1.617.2.4, UK lineage, from pango-designation issue #180 |
| AY.46 | 5 | Delta | Denmark 21.0%, Germany 11.0%, Sweden 8.0%, United States of America 7.0%, United Kingdom 6.0% | 2020-06-01 | Alias of B.1.617.2.46, Africa lineage, from pango-designation issue #247 |
| AY.16 | 5 | Delta | India 46.0%, United States of America 18.0%, **Kenya** 17.0%, Denmark 5.0%, Germany 3.0% | 2020-10-26 | Alias of B.1.617.2.16, lineage in **Kenya** and multiple other countries |
| AY.116 | 58 | Delta | Norway 12.0%, Sweden 11.0%, Brazil 10.0%, **Kenya** 8.0%, Zambia 8.0% | 2021-01-21 | Alias of B.1.617.2.116, Africa lineage |
| B.1.617.2 | 15 | Delta | United States of America 22.0%, India 19.0%, United Kingdom 13.0%, Turkey 7.0%, Germany 5.0% | 2021-03-01 | Predominantly India lineage with several spike mutations, pango-designation issue #49 |
| BA.1.1 | 14 | BA.1-like | United States of America 47.0%, United Kingdom 20.0%, Germany 7.0%, Canada 4.0%, France 2.0% | 2021-09-09 | Alias of B.1.1.529.1.1, from pango-designation issue #360 |
| BA.1.1.1 | 4 | BA.1-like | France 33.0%, Germany 18.0%, United Kingdom 11.0%, Spain 6.0%, Poland 5.0% | 2021-11-12 | Alias of B.1.1.529.1.1.1, European lineage |
| BA.1.9 | 1 | BA.1-like | Brazil 93.0%, United Kingdom 4.0%, United States of America 1.0%, Japan 0.0%, Chile 0.0% | 2021-12-11 | Alias of B.1.1.529.1.9, Brazil lineage |

| | | | | | |
|---|---|---|---|---|---|
| BA.2 | 1 | BA.2-like | United Kingdom 29.0%, Germany 13.0%, Denmark 12.0%, United States of America 11.0%, France 7.0% | 2021-09-21 | Alias of B.1.1.529.2, from pango-designation issue #361 |
| BA.2.31.1 | 4 | BA.2-like | Australia 28.0%, United States of America 27.0%, Uganda 17.0%, **Kenya** 9.0%, United Kingdom 3.0% | 2022-02-23 | Alias of B.1.1.529.2.31.1, mainly found in Uganda, defined by S:68T, from pango-designation issue #819 |
| BA.4.1 | 19 | BA.4-like | United States of America 48.0%, United Kingdom 9.0%, Germany 4.0%, Chile 4.0%, Israel 4.0% | 2021-12-14 | Alias of B.1.1.529.4.1, mainly found in South Africa, from pango-designation issue #548 |
| BA.4.1.9 | 2 | BA.4-like | United States of America 45.0%, South_Africa 7.0%, Denmark 6.0%, France 5.0%, Germany 5.0% | 2021-12-22 | Alias of B.1.1.529.4.1.9, mainly found in Zambia, USA, England and Mexico, from pango-designation issue #926 |
| BE.1 | 1 | BA.5-like | United States of America 28.0%, United Kingdom 15.0%, Germany 6.0%, Australia 5.0%, Japan 4.0% | 2020-07-21 | Alias of B.1.1.529.5.3.1.1, mainly found in South Africa, Austria and England, from pango-designation issue #625 |
| BA.5.2 | 3 | BA.5-like | United States of America 19.0%, Japan 17.0%, Germany 7.0%, United Kingdom 6.0%, France 4.0% | 2021-10-20 | Alias of B.1.1.529.5.2, mainly found in South Africa, England and USA, from pango-designation issue #551 |
| BA.5.2.1 | 41 | BA.5-like | United States of America 41.0%, Japan 9.0%, Canada 6.0%, Germany 5.0%, United Kingdom 5.0% | 2021-11-15 | Alias of B.1.1.529.5.2.1, mainly found in South Africa, England and USA, from pango-designation issue #657 |
| BA.5.2.20 | 1 | BA.5-like | United States of America 20.0%, France 11.0%, Germany 9.0%, United Kingdom 7.0%, Japan 7.0% | 2022-02-26 | Alias of B.1.1.529.5.2.20, mainly Indonesia, defined by C23707T on ORF1b:1050N branch to reduce unspecified BA.5.2 |
| BF.20 | 2 | BA.5-like | **Kenya** 28.0%, United Kingdom 18.0%, Canada 12.0%, United States of America 11.0%, Germany 8.0% | 2022-05-24 | Alias of B.1.1.529.5.2.1.20, lineage in **Kenya** and other countries |
| BF.9 | 3 | BA.5-like | Canada 64.0%, United States of America 32.0%, Germany 1.0%, Belgium 1.0%, United Kingdom 1.0% | 2022-05-28 | Alias of B.1.1.529.5.2.1.9, Canada lineage |
| BF.15 | 1 | BA.5-like | Australia 24.0%, United States of America 15.0%, United Kingdom 12.0%, Denmark 10.0%, Japan 6.0% | 2022-06-16 | Alias of B.1.1.529.5.2.1.15, Sri Lanka lineage, from pango-designation issue #893 |

**S2 Table.**

| Term | Definition |
|---|---|
| Household | A dwelling or a group of dwellings where the residents share the same kitchen or cooking space |
| *Index case* | The person with the earliest COVID-19-compatible symptoms onset date and laboratory confirmation (RT-PCR) of SARS-CoV-2 infection. |
| Stringency index (SI) | A measure based on nine government COVID-19 countermeasures response indicators rescaled to values between 0 and 100, with 100 being strictest (Hale et al., 2021). The nine response indicators used to form the SI are (1) school closures, (2) workplace closures, (3) cancellation of public events, (4) restrictions on public gatherings, (5) closures of public transport, (6) stay-at-home requirements, (7) public information campaigns, (8) restrictions on internal movements, and (9) international travel controls. |
| Multiple virus introduction | There are at least two viruses identified in a household during the follow-up period that appear to have been acquired outside the household, as indicated by genetic similarity with viruses isolated from the surrounding community. |
| Asymptomatic case | A study participant without current ARI symptoms at the time of collecting the sample or one week prior to that was found to carry the SARS-COV-2 genetic material. |

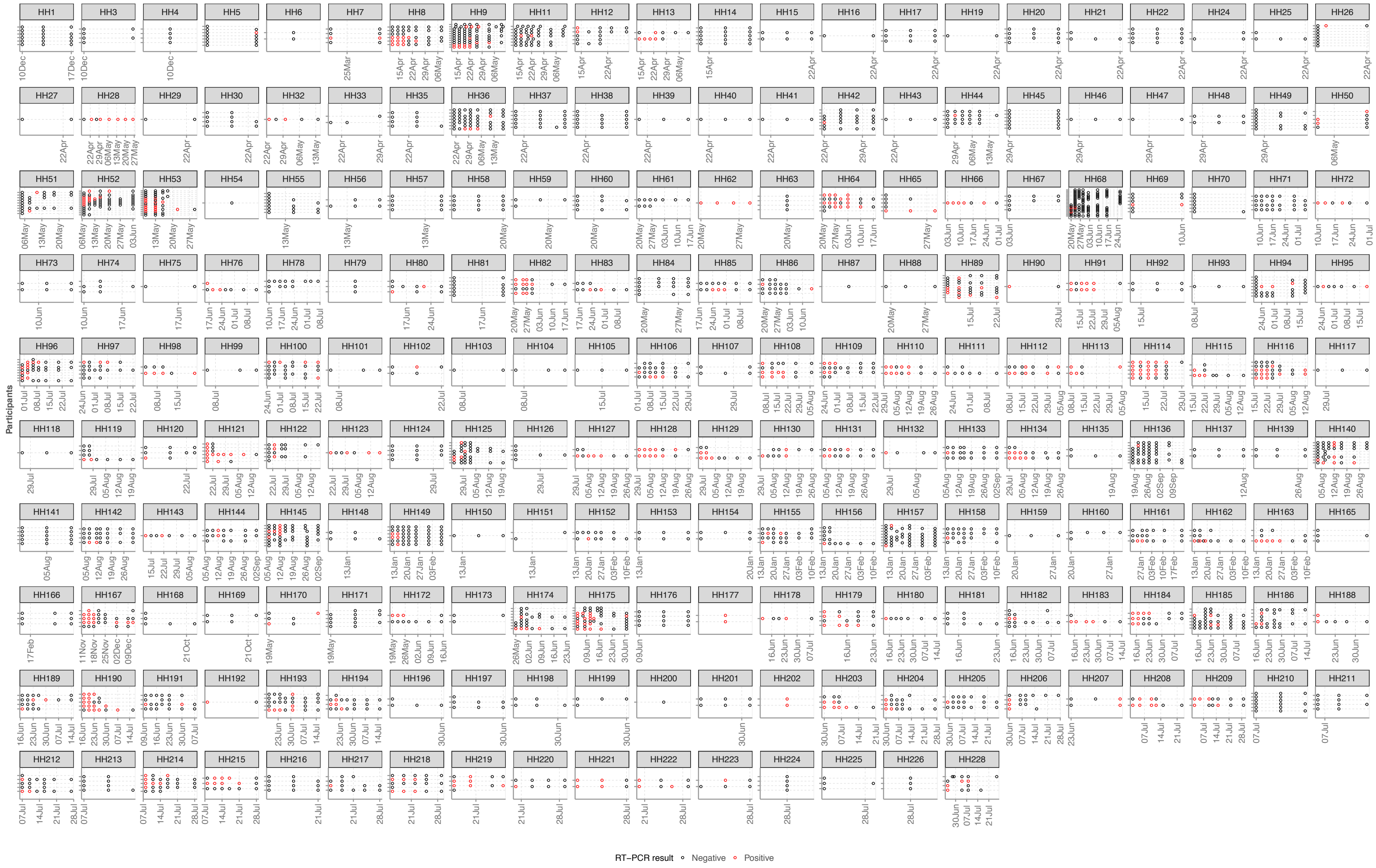

***S1 Figure. Sampling timelines and participant positivity in enrolled study households.*** *In total 214 households, 765 participants and 2780 NP/OP swabs were collected. A black circle refers to SARS-CoV-2 negative NP/OP, and a red-circle to a SARS-CoV-2 positive NP/OP.*

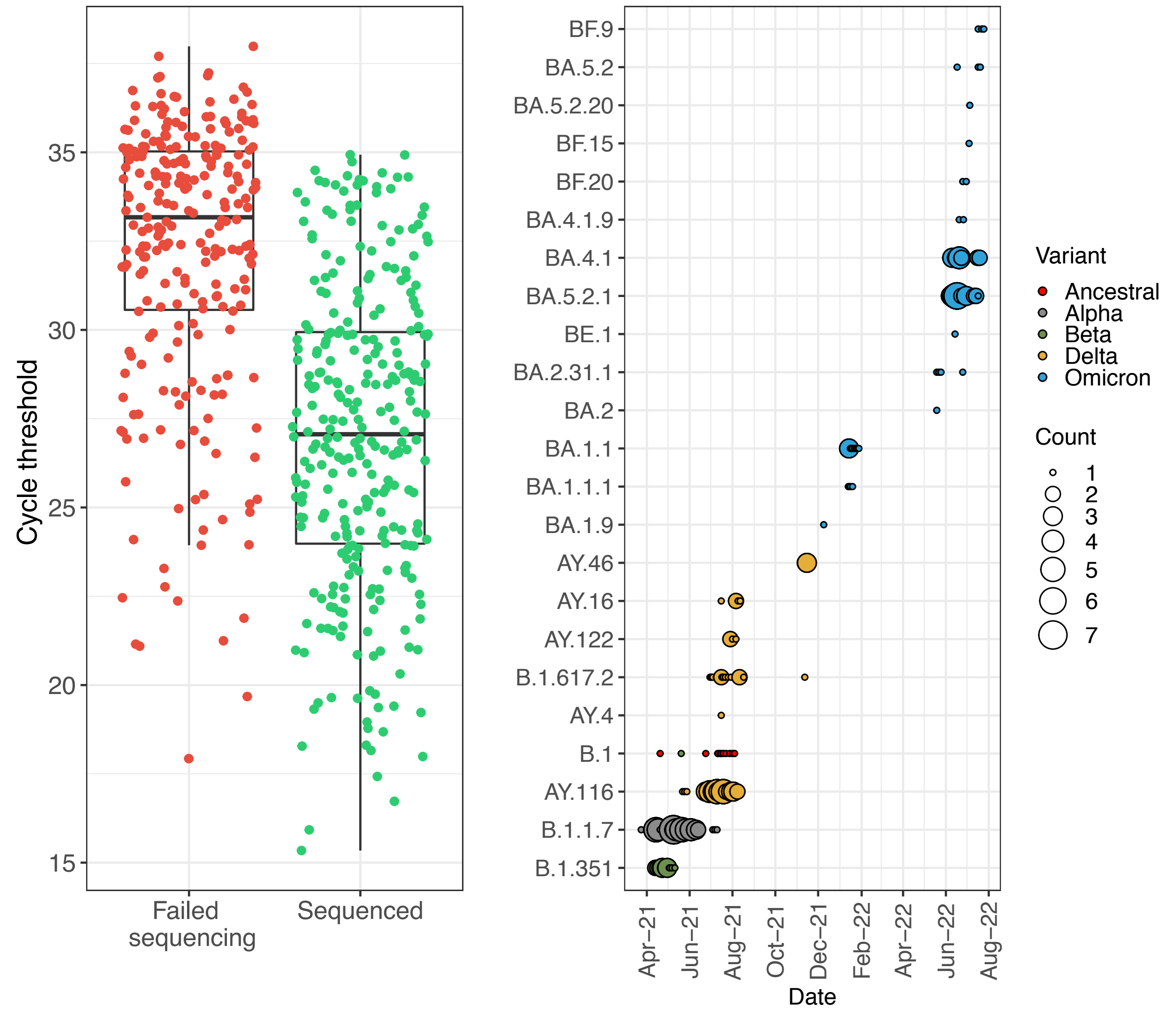

***S2 Figure. Study sequencing results summary.*** *Panel A: Comparison the cycle thresholds of failed versus successful sequenced RT-PCR positive NP/OP swabs. Panel B: Temporal distribution of lineages detected in the sequenced samples and their respective assignment into various variants of concern (VOC). Ancestral is represented by lineage B.1. The size of the circle is proportional to the number of genomes.*

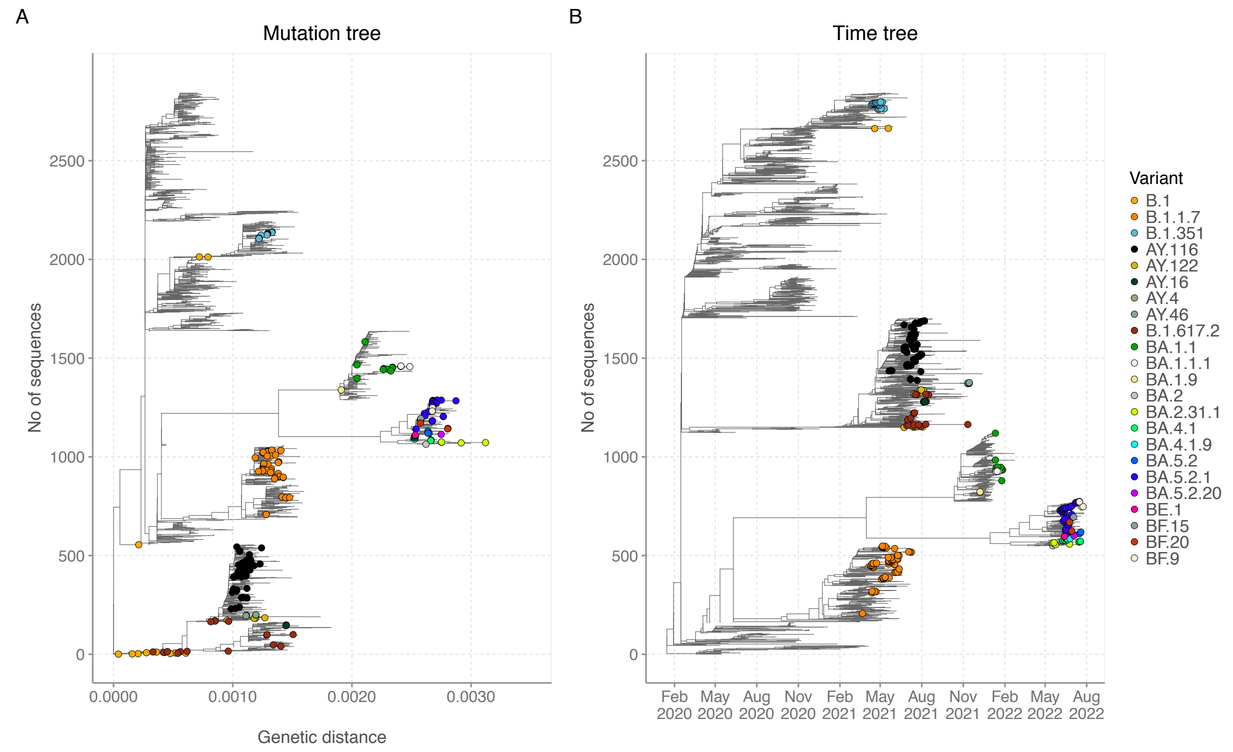

***S3 Figure. Phylogenetic relationships of the 2,843 SARS-CoV-2 consensus genomes analyzed in the present study.*** *These were gathered between 17$^{th}$ March 2020 and 26$^{th}$ July 2022 in coastal Kenya. The phylogenetic tree was obtained through the maximum likelihood phylogenetic inference. The resulting phylogeny includes 288 household study genomes (shown as filled circles at branch tips) and 2,555 other genomes (tips without symbols) sampled across the six Kenyan coastal counties. The household study genome tips are colored by assigned Pango lineage. We report both the original maximum likelihood tree (panel A, mid-point rooted) and a time-scaled version of it (panel B; see text for further detail).*

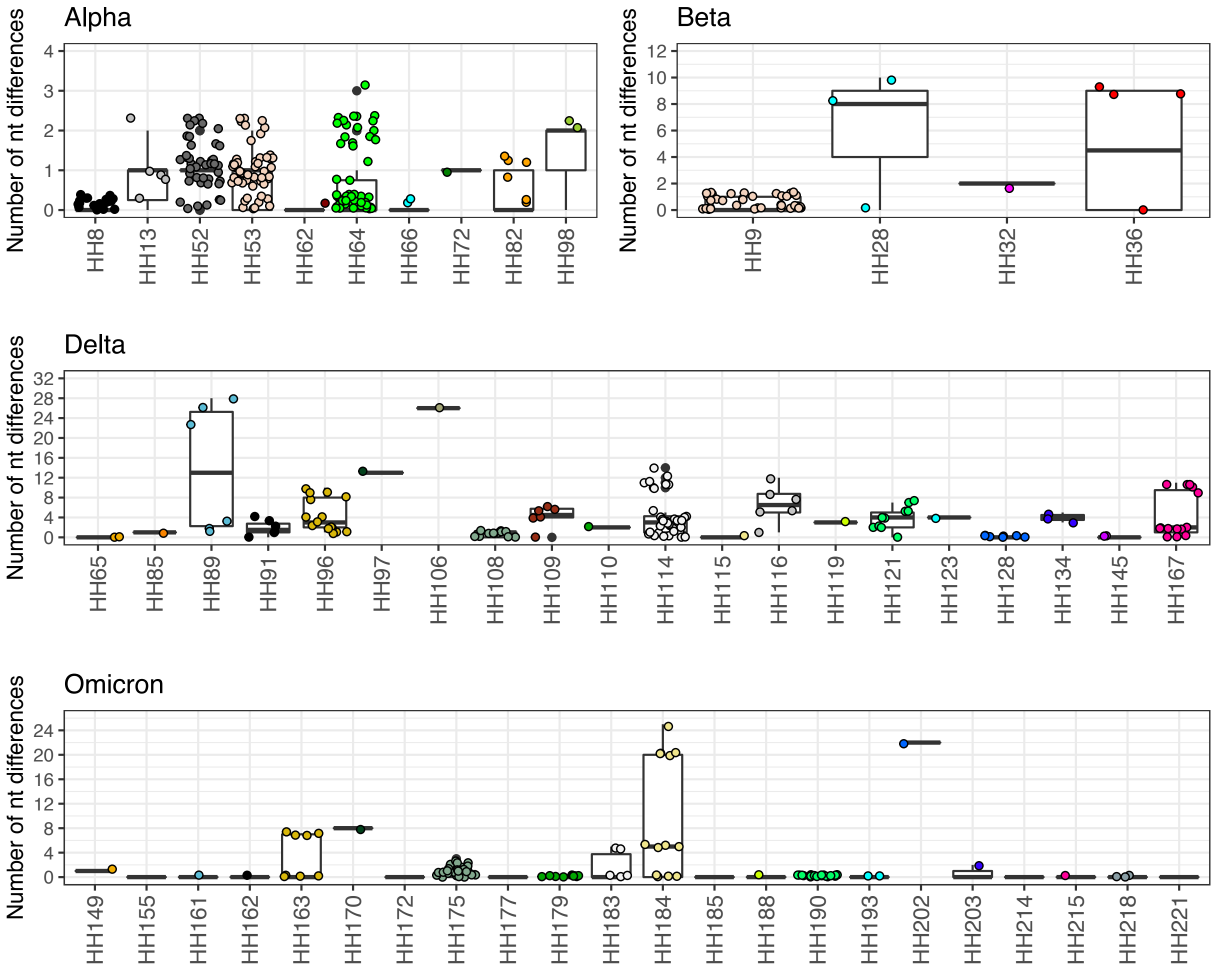

***S4 Figure. Comparison of pairwise nucleotide differences in the genome sequences of each household.*** *Panel A: Differences in the 10 Alpha infected households. Panel B: Differences in the four Beta infected households. Panel C: Differences in the 20 Delta infected households. Panel D: Differences in the 22 Omicron infected households. Households with a single genome sequence cannot been plotted.*

**Appendix:** Accession numbers of the genomic sequences from the household study deposited in the GISAID database

| GISAID name | GISAID accession number | HH_ID | Pango lineage |
|---|---|---|---|
| **hCoV-19/Kenya/C88814/2021** | EPI_ISL_6944234 | HH7 | B.1.1.7 |
| **hCoV-19/Kenya/C91180/2021** | EPI_ISL_2602534 | HH8 | B.1.1.7 |
| **hCoV-19/Kenya/C91759/2021** | EPI_ISL_2602552 | HH8 | B.1.1.7 |
| **hCoV-19/Kenya/C92331/2021** | EPI_ISL_2602577 | HH8 | B.1.1.7 |
| **hCoV-19/Kenya/C91760/2021** | EPI_ISL_2602553 | HH8 | B.1.1.7 |
| **hCoV-19/Kenya/C91181/2021** | EPI_ISL_2602535 | HH8 | B.1.1.7 |
| **hCoV-19/Kenya/C93346/2021** | EPI_ISL_2602602 | HH8 | B.1.1.7 |
| **hCoV-19/Kenya/C91761/2021** | EPI_ISL_2602554 | HH8 | B.1.1.7 |
| **hCoV-19/Kenya/C91182/2021** | EPI_ISL_2602536 | HH8 | B.1.1.7 |
| **hCoV-19/Kenya/C92333/2021** | EPI_ISL_2602578 | HH8 | B.1.1.7 |
| **hCoV-19/Kenya/C92271/2021** | EPI_ISL_3150137 | HH9 | B.1.351 |
| **hCoV-19/Kenya/C91600/2021** | EPI_ISL_2602546 | HH9 | B.1.351 |
| **hCoV-19/Kenya/C91601/2021** | EPI_ISL_2602547 | HH9 | B.1.351 |
| **hCoV-19/Kenya/C92272/2021** | EPI_ISL_3150138 | HH9 | B.1.351 |
| **hCoV-19/Kenya/C92937/2021** | EPI_ISL_2602588 | HH9 | B.1.351 |
| **hCoV-19/Kenya/C92585/2021** | EPI_ISL_2602582 | HH9 | B.1.351 |
| **hCoV-19/Kenya/C94719/2021** | EPI_ISL_7237336 | HH9 | B.1.351 |
| **hCoV-19/Kenya/C93639/2021** | EPI_ISL_2602615 | HH9 | B.1.351 |
| **hCoV-19/Kenya/C93186/2021** | EPI_ISL_2602594 | HH9 | B.1.351 |
| **hCoV-19/Kenya/C92733/2021** | EPI_ISL_7237332 | HH9 | B.1 |
| **hCoV-19/Kenya/C93763/2021** | EPI_ISL_2602616 | HH9 | B.1.351 |
| **hCoV-19/Kenya/C92283/2021** | EPI_ISL_3150139 | HH11 | B.1.1.7 |
| **hCoV-19/Kenya/C91774/2021** | EPI_ISL_2602555 | HH12 | B.1.1.7 |
| **hCoV-19/Kenya/C92336/2021** | EPI_ISL_2602579 | HH13 | B.1.1.7 |
| **hCoV-19/Kenya/C91765/2021** | EPI_ISL_2602570 | HH13 | B.1.1.7 |
| **hCoV-19/Kenya/C93179/2021** | EPI_ISL_6944188 | HH13 | B.1.1.7 |
| **hCoV-19/Kenya/C92757/2021** | EPI_ISL_2602583 | HH13 | B.1.1.7 |
| **hCoV-19/Kenya/C92339/2021** | EPI_ISL_2602580 | HH26 | B.1.351 |
| **hCoV-19/Kenya/C94433/2021** | EPI_ISL_2602632 | HH28 | B.1.351 |
| **hCoV-19/Kenya/C93185/2021** | EPI_ISL_2602593 | HH28 | B.1.351 |
| **hCoV-19/Kenya/C95523/2021** | EPI_ISL_3150140 | HH28 | B.1.351 |
| **hCoV-19/Kenya/C94432/2021** | EPI_ISL_2602631 | HH32 | B.1.351 |
| **hCoV-19/Kenya/C93178/2021** | EPI_ISL_2602612 | HH32 | B.1.351 |
| **hCoV-19/Kenya/C93778/2021** | EPI_ISL_2602603 | HH36 | B.1.351 |
| **hCoV-19/Kenya/C94434/2021** | EPI_ISL_2602633 | HH36 | B.1.351 |
| **hCoV-19/Kenya/C94773/2021** | EPI_ISL_6944125 | HH36 | B.1.351 |
| **hCoV-19/Kenya/C95899/2021** | EPI_ISL_2602678 | HH36 | B.1.351 |
| **hCoV-19/Kenya/C95162/2021** | EPI_ISL_2602641 | HH50 | B.1.1.7 |
| **hCoV-19/Kenya/C95530/2021** | EPI_ISL_3150141 | HH52 | B.1.1.7 |
| **hCoV-19/Kenya/C95531/2021** | EPI_ISL_3150142 | HH52 | B.1.1.7 |
| **hCoV-19/Kenya/C95837/2021** | EPI_ISL_2602674 | HH52 | B.1.1.7 |
| **hCoV-19/Kenya/C95532/2021** | EPI_ISL_3150143 | HH52 | B.1.1.7 |
| **hCoV-19/Kenya/C95838/2021** | EPI_ISL_2602675 | HH52 | B.1.1.7 |
| **hCoV-19/Kenya/C95533/2021** | EPI_ISL_2602669 | HH52 | B.1.1.7 |
| **hCoV-19/Kenya/C96392/2021** | EPI_ISL_3150148 | HH52 | B.1.1.7 |
| **hCoV-19/Kenya/C95534/2021** | EPI_ISL_3150144 | HH52 | B.1.1.7 |
| **hCoV-19/Kenya/C96568/2021** | EPI_ISL_2602701 | HH52 | B.1.1.7 |
| **hCoV-19/Kenya/C95839/2021** | EPI_ISL_2602676 | HH52 | B.1.1.7 |
| **hCoV-19/Kenya/C95564/2021** | EPI_ISL_6944206 | HH53 | B.1.1.7 |
| **hCoV-19/Kenya/C97616/2021** | EPI_ISL_5797402 | HH53 | B.1.1.7 |
| **hCoV-19/Kenya/C96381/2021** | EPI_ISL_3150145 | HH53 | B.1.1.7 |
| **hCoV-19/Kenya/C95565/2021** | EPI_ISL_2602670 | HH53 | B.1.1.7 |
| **hCoV-19/Kenya/C95566/2021** | EPI_ISL_2602671 | HH53 | B.1.1.7 |
| **hCoV-19/Kenya/C96388/2021** | EPI_ISL_3150147 | HH53 | B.1.1.7 |
| **hCoV-19/Kenya/C95567/2021** | EPI_ISL_2602672 | HH53 | B.1.1.7 |
| **hCoV-19/Kenya/C96386/2021** | EPI_ISL_3150146 | HH53 | B.1.1.7 |

| | | | |
|---|---|---|---|
| **hCoV-19/Kenya/C95569/2021** | EPI_ISL_2602722 | HH53 | B.1.1.7 |
| **hCoV-19/Kenya/C95570/2021** | EPI_ISL_2602723 | HH53 | B.1.1.7 |
| **hCoV-19/Kenya/C95571/2021** | EPI_ISL_2602724 | HH53 | B.1.1.7 |
| **hCoV-19/Kenya/C98175/2021** | EPI_ISL_2602742 | HH62 | B.1.1.7 |
| **hCoV-19/Kenya/C97303/2021** | EPI_ISL_2602721 | HH62 | B.1.1.7 |
| **hCoV-19/Kenya/C98887/2021** | EPI_ISL_5797413 | HH64 | B.1.1.7 |
| **hCoV-19/Kenya/C99753/2021** | EPI_ISL_3049440 | HH64 | B.1.1.7 |
| **hCoV-19/Kenya/C98636/2021** | EPI_ISL_5797411 | HH64 | B.1.1.7 |
| **hCoV-19/Kenya/C98223/2021** | EPI_ISL_2602746 | HH64 | B.1.1.7 |
| **hCoV-19/Kenya/C97554/2021** | EPI_ISL_2602732 | HH64 | B.1.1.7 |
| **hCoV-19/Kenya/C97866/2021** | EPI_ISL_2602737 | HH64 | B.1.1.7 |
| **hCoV-19/Kenya/C98222/2021** | EPI_ISL_2602745 | HH64 | B.1.1.7 |
| **hCoV-19/Kenya/C98886/2021** | EPI_ISL_7237388 | HH64 | B.1.1.7 |
| **hCoV-19/Kenya/C98888/2021** | EPI_ISL_6709405 | HH64 | B.1.1.7 |
| **hCoV-19/Kenya/C98635/2021** | EPI_ISL_7237375 | HH64 | B.1.1.7 |
| **hCoV-19/Kenya/C98225/2021** | EPI_ISL_2602747 | HH64 | B.1.1.7 |
| **hCoV-19/Kenya/C97868/2021** | EPI_ISL_2602738 | HH64 | B.1.1.7 |
| **hCoV-19/Kenya/C97556/2021** | EPI_ISL_5797400 | HH64 | B.1.1.7 |
| **hCoV-19/Kenya/C97622/2021** | EPI_ISL_2602734 | HH65 | AY.116 |
| **hCoV-19/Kenya/C97982/2021** | EPI_ISL_2602739 | HH65 | AY.116 |
| **hCoV-19/Kenya/C98497/2021** | EPI_ISL_2602748 | HH65 | AY.116 |
| **hCoV-19/Kenya/C99301/2021** | EPI_ISL_3049427 | HH66 | B.1.1.7 |
| **hCoV-19/Kenya/C98894/2021** | EPI_ISL_2602762 | HH66 | B.1.1.7 |
| **hCoV-19/Kenya/C99665/2021** | EPI_ISL_7237395 | HH66 | B.1.1.7 |
| **hCoV-19/Kenya/C100097/2021** | EPI_ISL_3049459 | HH66 | B.1.1.7 |
| **hCoV-19/Kenya/C97315/2021** | EPI_ISL_7237367 | HH68 | B.1.351 |
| **hCoV-19/Kenya/C99750/2021** | EPI_ISL_7237411 | HH69 | B.1.1.7 |
| **hCoV-19/Kenya/C100103/2021** | EPI_ISL_3049460 | HH72 | B.1.1.7 |
| **hCoV-19/Kenya/C99755/2021** | EPI_ISL_3049441 | HH72 | B.1.1.7 |
| **hCoV-19/Kenya/C100105/2021** | EPI_ISL_3049461 | HH80 | B.1.1.7 |
| **hCoV-19/Kenya/C97548/2021** | EPI_ISL_6709497 | HH82 | B.1.1.7 |
| **hCoV-19/Kenya/C98229/2021** | EPI_ISL_5797409 | HH82 | B.1.1.7 |
| **hCoV-19/Kenya/C97550/2021** | EPI_ISL_2602731 | HH82 | B.1.1.7 |
| **hCoV-19/Kenya/C97980/2021** | EPI_ISL_5797404 | HH82 | B.1.1.7 |
| **hCoV-19/Kenya/C97551/2021** | EPI_ISL_6709440 | HH82 | B.1.1.7 |
| **hCoV-19/Kenya/C101629/2021** | EPI_ISL_3049665 | HH85 | AY.116 |
| **hCoV-19/Kenya/C101330/2021** | EPI_ISL_3049550 | HH85 | AY.116 |
| **hCoV-19/Kenya/C103808/2021** | EPI_ISL_3150081 | HH89 | AY.116 |
| **hCoV-19/Kenya/C104088/2021** | EPI_ISL_7236562 | HH89 | B.1 |
| **hCoV-19/Kenya/C103610/2021** | EPI_ISL_3049852 | HH89 | AY.116 |
| **hCoV-19/Kenya/C103611/2021** | EPI_ISL_3049833 | HH89 | B.1.617.2 |
| **hCoV-19/Kenya/C103612/2021** | EPI_ISL_3049834 | HH89 | AY.116 |
| **hCoV-19/Kenya/C105933/2021** | EPI_ISL_7236630 | HH90 | B.1.617.2 |
| **hCoV-19/Kenya/C105699/2021** | EPI_ISL_3910032 | HH91 | AY.116 |
| **hCoV-19/Kenya/C103640/2021** | EPI_ISL_3150059 | HH91 | AY.116 |
| **hCoV-19/Kenya/C104665/2021** | EPI_ISL_7236567 | HH91 | B.1 |
| **hCoV-19/Kenya/C104879/2021** | EPI_ISL_6944143 | HH91 | AY.116 |
| **hCoV-19/Kenya/C105700/2021** | EPI_ISL_3910035 | HH91 | AY.116 |
| **hCoV-19/Kenya/C101329/2021** | EPI_ISL_7236506 | HH94 | B.1 |
| **hCoV-19/Kenya/C102007/2021** | EPI_ISL_3049691 | HH94 | AY.116 |
| **hCoV-19/Kenya/C103652/2021** | EPI_ISL_7236548 | HH94 | B.1 |
| **hCoV-19/Kenya/C101996/2021** | EPI_ISL_7236517 | HH96 | B.1.617.2 |
| **hCoV-19/Kenya/C101998/2021** | EPI_ISL_6944187 | HH96 | AY.116 |
| **hCoV-19/Kenya/C102000/2021** | EPI_ISL_7236518 | HH96 | AY.116 |
| **hCoV-19/Kenya/C102433/2021** | EPI_ISL_3049846 | HH96 | AY.116 |
| **hCoV-19/Kenya/C103609/2021** | EPI_ISL_6944136 | HH96 | AY.116 |
| **hCoV-19/Kenya/C102883/2021** | EPI_ISL_6944149 | HH96 | AY.116 |
| **hCoV-19/Kenya/C103225/2021** | EPI_ISL_3049850 | HH97 | AY.116 |
| **hCoV-19/Kenya/C102438/2021** | EPI_ISL_7236525 | HH97 | B.1.617.2 |
| **hCoV-19/Kenya/C103638/2021** | EPI_ISL_3150058 | HH98 | B.1.1.7 |
| **hCoV-19/Kenya/C102434/2021** | EPI_ISL_3049776 | HH98 | B.1.1.7 |
| **hCoV-19/Kenya/C103187/2021** | EPI_ISL_3049826 | HH98 | B.1.1.7 |

| | | | |
|---|---|---|---|
| **hCoV-19/Kenya/C104095/2021** | EPI_ISL_3150105 | HH100 | AY.116 |
| **hCoV-19/Kenya/C103648/2021** | EPI_ISL_3150064 | HH106 | AY.116 |
| **hCoV-19/Kenya/C102387/2021** | EPI_ISL_6944141 | HH106 | B.1.617.2 |
| **hCoV-19/Kenya/C103800/2021** | EPI_ISL_3150079 | HH108 | AY.116 |
| **hCoV-19/Kenya/C102873/2021** | EPI_ISL_3049797 | HH108 | AY.116 |
| **hCoV-19/Kenya/C103801/2021** | EPI_ISL_3150080 | HH108 | AY.116 |
| **hCoV-19/Kenya/C102874/2021** | EPI_ISL_3049798 | HH108 | AY.116 |
| **hCoV-19/Kenya/C104664/2021** | EPI_ISL_3150127 | HH108 | AY.116 |
| **hCoV-19/Kenya/C102872/2021** | EPI_ISL_3049796 | HH108 | AY.116 |
| **hCoV-19/Kenya/C101333/2021** | EPI_ISL_3049551 | HH109 | AY.116 |
| **hCoV-19/Kenya/C101335/2021** | EPI_ISL_3049552 | HH109 | AY.116 |
| **hCoV-19/Kenya/C102004/2021** | EPI_ISL_3049751 | HH109 | AY.116 |
| **hCoV-19/Kenya/C101636/2021** | EPI_ISL_3049666 | HH109 | AY.116 |
| **hCoV-19/Kenya/C106456/2021** | EPI_ISL_6944239 | HH110 | AY.116 |
| **hCoV-19/Kenya/C106338/2021** | EPI_ISL_3909866 | HH110 | AY.116 |
| **hCoV-19/Kenya/C103226/2021** | EPI_ISL_3049851 | HH112 | AY.116 |
| **hCoV-19/Kenya/C103642/2021** | EPI_ISL_3150060 | HH114 | AY.116 |
| **hCoV-19/Kenya/C103643/2021** | EPI_ISL_3150061 | HH114 | AY.116 |
| **hCoV-19/Kenya/C104668/2021** | EPI_ISL_7236571 | HH114 | B.1.617.2 |
| **hCoV-19/Kenya/C104886/2021** | EPI_ISL_7236600 | HH114 | B.1 |
| **hCoV-19/Kenya/C103644/2021** | EPI_ISL_3150062 | HH114 | AY.116 |
| **hCoV-19/Kenya/C103848/2021** | EPI_ISL_3150086 | HH114 | AY.116 |
| **hCoV-19/Kenya/C103850/2021** | EPI_ISL_6944204 | HH114 | AY.116 |
| **hCoV-19/Kenya/C104671/2021** | EPI_ISL_6944140 | HH114 | B.1.617.2 |
| **hCoV-19/Kenya/C103646/2021** | EPI_ISL_3150063 | HH114 | AY.116 |
| **hCoV-19/Kenya/C104887/2021** | EPI_ISL_6944213 | HH114 | AY.116 |
| **hCoV-19/Kenya/C104672/2021** | EPI_ISL_3150128 | HH115 | B.1.617.2 |
| **hCoV-19/Kenya/C104673/2021** | EPI_ISL_3150129 | HH115 | AY.16 |
| **hCoV-19/Kenya/C104674/2021** | EPI_ISL_3150130 | HH116 | AY.116 |
| **hCoV-19/Kenya/C104676/2021** | EPI_ISL_3150131 | HH116 | AY.116 |
| **hCoV-19/Kenya/C105216/2021** | EPI_ISL_6709422 | HH116 | B.1.617.2 |
| **hCoV-19/Kenya/C104873/2021** | EPI_ISL_6944124 | HH116 | AY.116 |
| **hCoV-19/Kenya/C106194/2021** | EPI_ISL_3910162 | HH119 | AY.116 |
| **hCoV-19/Kenya/C105748/2021** | EPI_ISL_7236622 | HH119 | AY.116 |
| **hCoV-19/Kenya/C104727/2021** | EPI_ISL_7236591 | HH120 | B.1.617.2 |
| **hCoV-19/Kenya/C104889/2021** | EPI_ISL_6944123 | HH121 | AY.116 |
| **hCoV-19/Kenya/C105765/2021** | EPI_ISL_6944205 | HH121 | AY.116 |
| **hCoV-19/Kenya/C106395/2021** | EPI_ISL_7236665 | HH121 | B.1 |
| **hCoV-19/Kenya/C106189/2021** | EPI_ISL_3910159 | HH121 | AY.116 |
| **hCoV-19/Kenya/C104890/2021** | EPI_ISL_7236611 | HH121 | B.1.617.2 |
| **hCoV-19/Kenya/C104892/2021** | EPI_ISL_6944192 | HH121 | AY.116 |
| **hCoV-19/Kenya/C105691/2021** | EPI_ISL_7236621 | HH122 | B.1 |
| **hCoV-19/Kenya/C107193/2021** | EPI_ISL_7236708 | HH123 | B.1.617.2 |
| **hCoV-19/Kenya/C106832/2021** | EPI_ISL_7236700 | HH123 | B.1.617.2 |
| **hCoV-19/Kenya/C106324/2021** | EPI_ISL_7236651 | HH127 | B.1.617.2 |
| **hCoV-19/Kenya/C106334/2021** | EPI_ISL_3909859 | HH128 | AY.116 |
| **hCoV-19/Kenya/C106458/2021** | EPI_ISL_3909896 | HH128 | AY.116 |
| **hCoV-19/Kenya/C106697/2021** | EPI_ISL_3909947 | HH128 | AY.116 |
| **hCoV-19/Kenya/C106335/2021** | EPI_ISL_3909862 | HH128 | AY.116 |
| **hCoV-19/Kenya/C106589/2021** | EPI_ISL_3909920 | HH128 | AY.116 |
| **hCoV-19/Kenya/C106459/2021** | EPI_ISL_3909900 | HH128 | AY.116 |
| **hCoV-19/Kenya/C106696/2021** | EPI_ISL_3909942 | HH128 | AY.116 |
| **hCoV-19/Kenya/C106181/2021** | EPI_ISL_7236638 | HH129 | B.1 |
| **hCoV-19/Kenya/C106386/2021** | EPI_ISL_3909872 | HH129 | AY.116 |
| **hCoV-19/Kenya/C106630/2021** | EPI_ISL_3909936 | HH133 | AY.122 |
| **hCoV-19/Kenya/C106262/2021** | EPI_ISL_3910200 | HH134 | AY.122 |
| **hCoV-19/Kenya/C106540/2021** | EPI_ISL_7236687 | HH134 | B.1 |
| **hCoV-19/Kenya/C106397/2021** | EPI_ISL_3909875 | HH134 | AY.122 |
| **hCoV-19/Kenya/C106263/2021** | EPI_ISL_6944157 | HH134 | AY.122 |
| **hCoV-19/Kenya/C106828/2021** | EPI_ISL_7236695 | HH142 | B.1.617.2 |
| **hCoV-19/Kenya/C103805/2021** | EPI_ISL_7236554 | HH143 | B.1 |
| **hCoV-19/Kenya/C113902/2022** | EPI_ISL_11116830 | HH149 | BA.1.1 |

| | | | |
|---|---|---|---|
| **hCoV-19/Kenya/C113903/2022** | EPI_ISL_11116831 | HH149 | BA.1.1 |
| **hCoV-19/Kenya/C113981/2022** | EPI_ISL_11116843 | HH155 | BA.1.1 |
| **hCoV-19/Kenya/C114051/2022** | EPI_ISL_11116848 | HH155 | BA.1.1 |
| **hCoV-19/Kenya/C114287/2022** | EPI_ISL_11116859 | HH155 | BA.1.1 |
| **hCoV-19/Kenya/C113966/2022** | EPI_ISL_11116840 | HH156 | BA.1.1.1 |
| **hCoV-19/Kenya/C113980/2022** | EPI_ISL_11116842 | HH157 | BA.1.1 |
| **hCoV-19/Kenya/C113972/2022** | EPI_ISL_11116841 | HH158 | BA.1.1 |
| **hCoV-19/Kenya/C114317/2022** | EPI_ISL_11116862 | HH161 | BA.1.1 |
| **hCoV-19/Kenya/C114418/2022** | EPI_ISL_11116864 | HH161 | BA.1.1 |
| **hCoV-19/Kenya/C113936/2022** | EPI_ISL_11116838 | HH162 | BA.1.1.1 |
| **hCoV-19/Kenya/C114136/2022** | EPI_ISL_11116853 | HH162 | BA.1.1.1 |
| **hCoV-19/Kenya/C114017/2022** | EPI_ISL_11116846 | HH162 | BA.1.1.1 |
| **hCoV-19/Kenya/C114222/2022** | EPI_ISL_11116857 | HH163 | BA.1.1 |
| **hCoV-19/Kenya/C114133/2022** | EPI_ISL_11116852 | HH163 | BA.1.1 |
| **hCoV-19/Kenya/C113933/2022** | EPI_ISL_11116837 | HH163 | BA.1.1 |
| **hCoV-19/Kenya/C114020/2022** | EPI_ISL_11116847 | HH163 | BA.1.1 |
| **hCoV-19/Kenya/C114359/2022** | EPI_ISL_11116863 | HH163 | BA.1.1 |
| **hCoV-19/Kenya/C111708/2021** | EPI_ISL_6709504 | HH167 | AY.46 |
| **hCoV-19/Kenya/C112904/2021** | EPI_ISL_11116699 | HH167 | BA.1.9 |
| **hCoV-19/Kenya/C111861/2021** | EPI_ISL_6944217 | HH167 | AY.46 |
| **hCoV-19/Kenya/C111709/2021** | EPI_ISL_6709400 | HH167 | B.1.617.2 |
| **hCoV-19/Kenya/C111863/2021** | EPI_ISL_6944218 | HH167 | AY.46 |
